# Graph-Based Clinical Recommender: Predicting Specialists Procedure Orders using Graph Representation Learning

**DOI:** 10.1101/2022.11.21.22282571

**Authors:** Sajjad Fouladvand, Federico Reyes Gomez, Hamed Nilforoshan, Matthew Schwede, Morteza Noshad, Olivia Jee, Jiaxuan You, Rok Sosic, Jure Leskovec, Jonathan Chen

**Author notes:** Corresponding Author: Sajjad Fouladvand, 3180 Porter Drive, Room A223.11, Palo Alto 94306, CA, Twitter: @SajjadFV. Authors contributed equally to this manuscript (Sajjad Fouladvand, Federico Reyes Gomez).

## Abstract

**Objective:** To determine whether graph neural network based models of electronic health records can predict specialty consultation care needs for endocrinology and hematology more accurately than the standard of care checklists and other conventional medical recommendation algorithms in the literature.

**Methods:** Demand for medical expertise far outstrips supply, with tens of millions in the US alone with deficient access to specialty care. Rather than potentially months long delays to initiate diagnostic workup and medical treatment with a specialist, referring primary care supported by an automated recommender algorithm could anticipate and directly initiate patient evaluation that would otherwise be needed at subsequent a specialist appointment. We propose a novel graph representation learning approach with a heterogeneous graph neural network to model structured electronic health records and formulate recommendation/prediction of subsequent specialist orders as a link prediction problem.

**Results:** Models are trained and assessed in two specialty care sites: endocrinology and hematology. Our experimental results show that our model achieves an 8% improvement in ROC-AUC for endocrinology (ROC-AUC=0.88) and 5% improvement for hematology (ROC-AUC=0.84) personalized procedure recommendations over prior medical recommender systems. These recommender algorithm approaches provide medical procedure recommendations for endocrinology referrals more effectively than manual clinical checklists (recommender: precision=0.60, recall=0.27, F1-score=0.37) vs. (checklist: precision=0.16, recall=0.28, F1-score=0.20), and similarly for hematology referrals (recommender: precision=0.44, recall=0.38, F1-score=0.41) vs. (checklist: precision=0.27, recall=0.71, F1-score=0.39).

**Conclusion:** Embedding graph neural network models into clinical care can improve digital specialty consultation systems and expand the access to medical experience of prior similar cases.

## 1. Introduction

Access to medical specialty care is often delayed due to growing limitations in clinicians’ time and resources leading to over 25 million Americans with deficient access to specialty care (Woolhandler and Himmelstein, 2017), associated with higher mortality (Prentice and Pizer, 2007). Prediction of medical procedures to be ordered during initial outpatient specialty consultation care can facilitate specialist consultations by eliminating delay and further follow-up of diagnostic steps, in some cases completely eliminating the need for an in-person consultation (Chiang et al., 2020; Kim-Hwang et al., 2010). Clinical checklists are the current standard of practice to improve specialty referral healthcare delivery (Fantasia et al., 2021; Keely et al., 2013; Siepierski, 2013; Vimalananda et al., 2015) and in critical situations such as those resulting from the COVID-19 pandemic (Webster, 2020).

Clinical checklists are labor-intensive to manually produce while both being not easily generalizable or personalizable to complex scenarios when they largely offer one-size-fits-all generic guidance and checklists for considerations (Kumar et al., 2020; Tricoci et al., 2009; Li et al., 2019; McGlynn et al., 2003). Automated AI systems could improve specialty care systems by providing personalized recommendations based on prior subspecialist care, addressing limitations in general guidelines and checklists (Middleton et al., 2016; Berner and La Lande, 2016; Bright et al., 2012). Leveraging artificial intelligence (AI) models trained using large scale routinely collected electronic health records (EHR) to create automatic specialty care procedure recommendation could improve efficient use of scarce clinician time and in turn increase access for more patients to reach appropriate care and consultation.

To this end, there have been multiple attempts to create automated and data-driven medical order recommender systems. OrderRex (Chen et al., 2016) was created based on association statistics and Bayesian rules to show promising results in improving clinical order decision making process and usability (Kumar et al., 2020). Ip et al. (2022) used co-occurrence statistics to create a recommender algorithm to predict pediatric endocrinology patients’ initial workup needs. Classical machine learning models (Hunter-Zinck et al., 2019), assessing coverage of manually authored order sets using optimization-based and clustering techniques (Zhang et al., 2014), item-based collaborative filtering (Klann et al., 2009), and artificial neural networks were used for personalized general clinical orders (Wang et al., 2020) and endocrinology procedure recommendation (Noshad et al., 2021).

To improve upon prior methods, we consider that the heterogeneity and structured nature of electronic health records (EHR) can be captured more effectively using graphical models (Park et al., 2022; Choi et al., 2018, 2017). A Graph Convolutional Transformer (GCT) (Choi et al., 2020) maps encounters into a fully connected graph and infers the underlying structure by computing self-attentions on the graph connection. Liu et al. (2020) addressed the high visibility (Li et al., 2018) of hub nodes such as demographic nodes and showed the effectiveness of modeling EHR data into heterogeneous graphs. Further, heterogeneous graph neural networks (GNN) have been utilized in drug pair side effect prediction (Zitnik et al., 2018), medical diagnosis prediction (Liu et al., 2021) and medical concept representations (Wu et al., 2021; Vretinaris et al., 2021).

Motivated by Hamilton et al. (2017), Zitnik et al. (2018), and Veličković et al. (2018) we propose a novel GNN-based framework to provide personalized procedure order recommendations prior to or during patients’ initial specialty care visits. Note, here we use the terminology ‘order’ to refer to the procedures ordered by physicians (e.g., laboratory tests, imaging studies, additional referrals and consultations).

We tested our models on Endocrinology and Hematology specialty referral care as two of the most common specialties that are receptive to virtual consultations given their strong basis in structured diagnostic test results (Palen et al., 2012; Keely et al., 2013; Liddy et al., 2013). Our objective is to determine whether GNN based models of EHR data can predict specialty consultation care needs for endocrine and hematology more accurately than the standard of care guidelines and checklists and other conventional medical recommendation algorithms in the literature.

## 2. Materials and Methods

Figure 1 shows the overall schema of our proposed framework. We mapped patients’ historical EHR data recorded prior to the patients’ first referrals to specialty care clinics into a heterogeneous graph neural network. This model was trained to predict procedures ordered by endocrinology and hematology specialists during patients visits at the specialty care clinics.

**Figure 1:**
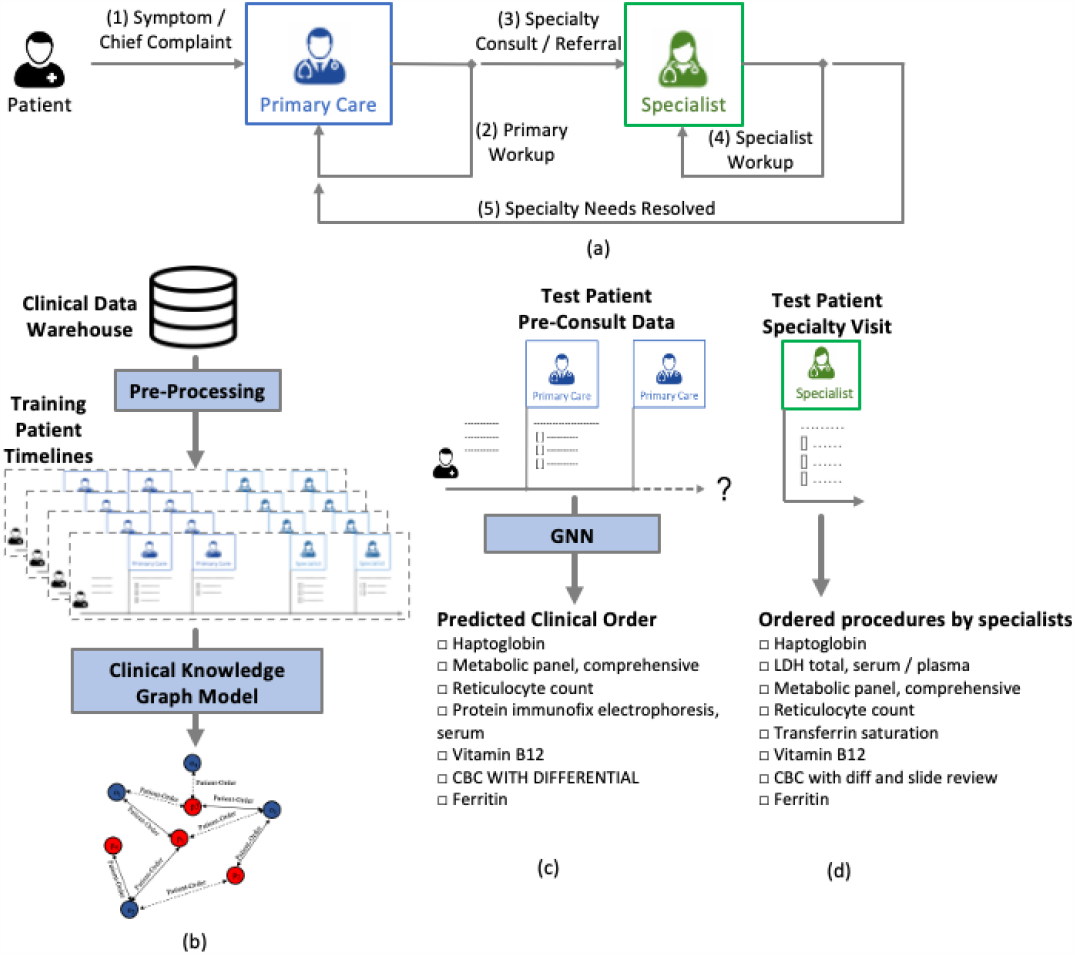
Overall schema of the proposed framework. (a) Specialists’ orders and workup can be initiated by primary care providers at referral or consultation time. (b) Patients’ historical EHR data including diagnoses, procedures, and lab results were used to create a heterogeneous GNN. Nodes are patients and orders. Solid line edges show orders before referral and dashed line edges show the specialists orders after referral date. (c) The GNN model was used to predict future specialists’ orders. This figure shows the GNN model recommendations for one example patient referred to the hematology clinic. (d) Procedures ordered by specialists (ground truth) for the example patient shown in (c). This list highly overlaps with the procedures recommended by the GNN model (c).

### 2.1. Endocrinology Data

Our data includes all outpatients referred by XXXX primary care providers to the XXXX Endocrinology clinic between January 2008 and December 2018. Use of this data for this study was approved by XXXX Institutional Review Board. We only included patients’ first visit with the respective specialist within four months of their referral dates to reflect initial engagement with the specialist in response to to the referral consultation request. Our final data set include 6,821 new referrals to the endocrinology clinic.

We denote the list of patient referrals as ***P*** = {*p*_1_, …, *p*_*n*_} in which *n* is the number of patient referrals. Each patient referral *p*_*i*_ constitutes a tuple (*t*_*i*_, ***D***^*i*^, ***O***^*i*^, ***L***^*i*^, ***Y*** ^*i*^), where *t*_*i*_ is referral’s date and ***D***^*i*^ ∈ ℝ^10^, ***O***^*i*^ ∈ ℝ^60^, and ***L***^*i*^ ∈ ℝ^300×3^ are multi-hot encoded vectors representing diagnoses codes, procedure orders, and lab results for *p*_*i*_ recorded prior to *t*_*i*_. We used a two month look back window for lab results and procedures. Each lab result was converted to a vector with three elements indicating (a) if *p*_*i*_ has had the lab result, (b) if the result was high, and (c) if the result was low. ***Y***^*i*^ is a multi-hot encoded vector representing the procedures ordered by the specialist during patient’s special care visit. Our final feature set includes 370 features. The target set includes 60 procedure orders. A full list of diagnoses, procedures and lab tests are presented in Table A1, Table A2, and Table A3 in Appendix A, respectively.

### 2.2. Hematology Data

Our hematology cohort includes all outpatients referred to XXXX Hematology clinic by primary care providers as new patients from 2008 to 2021. Our final cohort includes 2,007 patients. Data format is similar to the Endocrinology cohort described in section 2.1. Feature set includes the top-100 most commonly recorded diagnoses, top-100 most commonly ordered labs as well as 33 medical procedures. The procedures were selected based on the health system’s internally produced checklists for clinic referrals and electronic consultations. Lab tests were one-hot encoded based on their results and flagged as one of the following options: abnormal, normal, low, low off-scale, low panic, high, high off-scale, high panic, negative, and positive. Each Hematology patient referral *p*_*i*_ constitute (*t*_*i*_, ***D***^*i*^, ***O***^*i*^, ***L***^*i*^, ***Y*** ^*i*^), where *t*_*i*_ is referral’s date and ***D***^*i*^ ∈ ℝ^100^, ***O***^*i*^ ∈ ℝ^33^, and ***L***^*i*^ ∈ ℝ^298×3^ are multi-hot encoded vectors representing diagnoses codes, procedure orders, and lab results for *p*_*i*_ prior to *t*_*i*_. Table A4 in Appendix A lists the diagnoses, Table A5 shows the lab tests, and Table A6 shows the procedures used in this study for the Hematology cohort.

### 2.3. Descriptive Analysis

Table 1 shows the cohort demographics. In general, demographics in terms of age, sex, race, and ethnicity were similar among endocrinology and hematology patients.

**Table 1:** Patient characteristics among endocrinology and hematology cohorts. Numbers are N(25^*th*^ percentile, 75^*th*^ percentile) for age and N(%) for other variables.

| Variable | Endocrinology<br>(n=6,802) | Hematology<br>(n=2007) |
| --- | --- | --- |
| Age | 52.85 (39, 66) | 58.93 (46, 72) |
| Female | 4,104 (66%) | 1,066 (53%) |
| **Race** |  |  |
| White | 3,098 (50%) | 1,131 (56%) |
| Other | 1,083 (17%) | 308 (15%) |
| Asian | 1,420 (23%) | 381 (19%) |
| Black | 287 (5%) | 122 (6%) |
| Unknown | 245 (4%) | 21 (2%) |
| Pacific Islander | 82 (1%) | 25 (1%) |
| Native American | 23 (0.4%) | 8 (0.4%) |
| **Ethnicity** |  |  |
| Non Hisp./Lat. | 5,251 (84%) | 1,738 (86%) |
| Hisp./Lat. | 737 (12%) | 234 (12%) |
| Unknown | 250 (4%) | 35 (2%) |

Tables 2 and 3 show the most common diagnoses, procedures and medications recorded for endocrinology and hematology referrals before patients’ referral dates, respectively. Variable frequencies for each cohort were computed against new patients to all other specialty care clinics except our study cohorts. For our endocrinology cohort, the top-3 most frequently observed diagnosis codes were thyroid nodule, hyperthyroidism and hypothyroidism, and the top-3 most frequently ordered procedures were comprehensive metabolic panel, thyroid-stimulating hormone test (TSH), and T4 free. The Top-3 most frequently prescribed medications for this cohort were ondansetron, acetaminophen, and Normal Saline IV Bolus (reflecting supportive treatments that accompany specialty treatments like chemotherapy).

**Table 2:**
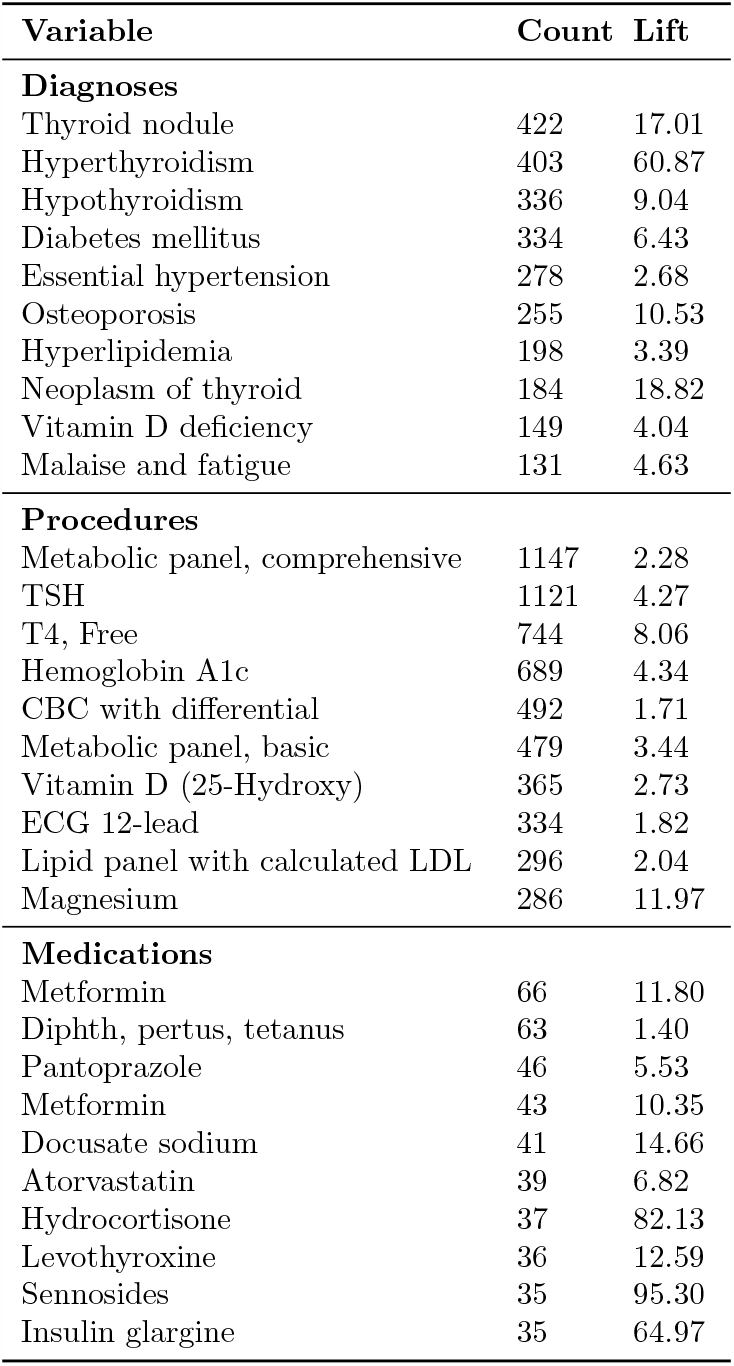
Top-10 most observed diagnoses, procedures, and medications in endocrinology patients records. Count shows the number of encounters and Lift shows the ratio of frequency of each variable in the endocrinology cohort to its frequency in all specialty clinics except endocrinology.

| Variable | Count | Lift |
| --- | --- | --- |
| **Diagnoses** |  |  |
| Thyroid nodule | 422 | 17.01 |
| Hyperthyroidism | 403 | 60.87 |
| Hypothyroidism | 336 | 9.04 |
| Diabetes mellitus | 334 | 6.43 |
| Essential hypertension | 278 | 2.68 |
| Osteoporosis | 255 | 10.53 |
| Hyperlipidemia | 198 | 3.39 |
| Neoplasm of thyroid | 184 | 18.82 |
| Vitamin D deficiency | 149 | 4.04 |
| Malaise and fatigue | 131 | 4.63 |
| **Procedures** |  |  |
| Metabolic panel, comprehensive | 1147 | 2.28 |
| TSH | 1121 | 4.27 |
| T4, Free | 744 | 8.06 |
| Hemoglobin A1c | 689 | 4.34 |
| CBC with differential | 492 | 1.71 |
| Metabolic panel, basic | 479 | 3.44 |
| Vitamin D (25-Hydroxy) | 365 | 2.73 |
| ECG 12-lead | 334 | 1.82 |
| Lipid panel with calculated LDL | 296 | 2.04 |
| Magnesium | 286 | 11.97 |
| **Medications** |  |  |
| Metformin | 66 | 11.80 |
| Diphth, pertus, tetanus | 63 | 1.40 |
| Pantoprazole | 46 | 5.53 |
| Metformin | 43 | 10.35 |
| Docusate sodium | 41 | 14.66 |
| Atorvastatin | 39 | 6.82 |
| Hydrocortisone | 37 | 82.13 |
| Levothyroxine | 36 | 12.59 |
| Sennosides | 35 | 95.30 |
| Insulin glargine | 35 | 64.97 |

**Table 3:** Top-10 most observed diagnoses, procedures, and medications in hematology patients records. Count shows the number of encounters and Lift shows the ratio of frequency of each variable in the hematology cohort to its frequency in all specialty clinics except hematology.

| Variable | Count | Lift |
| --- | --- | --- |
| **Diagnoses** |  |  |
| Anemia | 365 | 37.66 |
| Thrombocytopenia | 345 | 45.92 |
| Essential hypertension | 200 | 2.7 |
| Hyperlipidemia | 147 | 3.51 |
| Iron deficiency anemia | 139 | 21.47 |
| Pulmonary embolism and infarction | 128 | 53.25 |
| Leukopenia | 118 | 75.44 |
| Deep venous thrombosis of lower extremity | 103 | 34.79 |
| Diabetes mellitus | 101 | 2.54 |
| Shortness of breath | 101 | 4.39 |
| **Procedures** |  |  |
| Metabolic panel, comprehensive | 1,353 | 2.42 |
| CBC with differential | 937 | 2.6 |
| Prothrombin time | 825 | 8.79 |
| ECG 12-lead | 722 | 3.89 |
| PTT partial thromboplastin time | 614 | 13.31 |
| Ferritin | 580 | 8.2 |
| Lactate Dehydrogenase (LDH) | 567 | 14.89 |
| Up ad lib | 551 | 4143.86 |
| Magnesium | 549 | 22.46 |
| Sequential compression device (SCD) | 519 | 8.96 |
| **Medications** |  |  |
| Acyclovir | 120 | 39.46 |
| Pantoprazole | 118 | 12.74 |
| Polyethylene glycol | 114 | 36.90 |
| Sennosides | 89 | 157.00 |
| Docusate sodium | 78 | 27.62 |
| Acetaminophen | 73 | 152.28 |
| Oxycodone | 72 | 12.33 |
| Sulfamethoxazole-trimethoprim | 70 | 7.95 |
| Apixaban | 61 | 46.12 |
| Ferrous sulfate | 58 | 37.08 |

Anemia, thrombocytopenia and essential hypertension are the top-3 most frequently observed diagnosis codes recorded for hematology patients. Comprehensive metabolic panel, CBC, prothrombin time were most frequently ordered procedures and ondansetron, acetaminophen and polyethylene glycol were the most frequently prescribed medications for hematology patients. Table 3 shows the full list of the top-10 most frequently observed diagnoses, procedures and medications in hematology patients.

### 2.4. Proposed Method

#### 2.4.1. Graph Structure

We modeled patients’ EHR data into a heterogeneous graph neural network *G* = (*V, E*) (see Figure 2(*a*)). *V* contains two node types: patient referral nodes 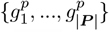, and procedure order nodes 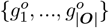. Each patient node 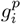 is assigned a multi-hot encoded feature vector consisting of the concatenation of ***D***^*i*^ and ***L***^*i*^ and each procedure order node 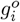 are associated with one-hot encoding of the entity IDs (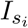 and 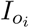, respectively).

**Figure 2:**
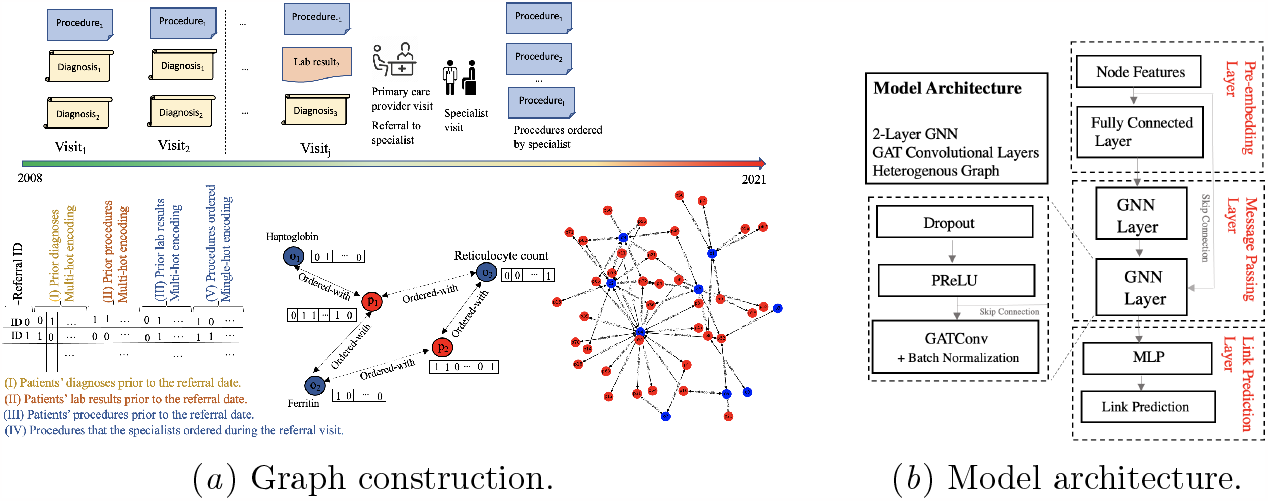
Patients EHR data are formatted in a multi-hot encoding matrix. Red nodes show patients and blue nodes show procedures. Each patient node is assigned with a multi-hot encoded feature vector consisting of the concatenation of diagnoses and lab results features, and each procedure order node are associated with one-hot encoding of the procedure IDs. Solid edges show procedures ordered before referral dates and dashed edges show procedures ordered by specialists and after referral dates (targets).

Edge set *E* contains two edge types. ‘ordered-with’ edges with edge labels set to 0 that are edges between patient nodes and the procedures they have done before *t*_*i*_, and ‘ordered-with’ edges with edge label set to 1 that connect the patients with the procedures that their specialist ordered during the specialty care visit after *t*_*i*_. Note, ‘ordered-with’ edges with edge labels equal to 1 that represent specialist’s orders after referral date were not used during training and were only used in the prediction phase as we are aiming to predict procedure orders after *t*_*i*_. We formulate this task as binary link prediction of the existence of ‘ordered-with’ edges between a patient and an order. Further, node degree, node clustering coefficient and centrality transformations were applied to add synthetic features to each node feature vector. While the model can learn those features on its own, we added them to help the model focus on learning other features. We apply a different graph convolutional layers with independent parameters to each message type of (head, relation, tail) and aggregate embeddings across all node types. The same graph attention mechanism was applied to all node types.

#### 2.4.2. Message Passing and Graph Attention

Figure 2(*b*) shows our proposed architecture. A fully connected layer with hidden size of 128 was used to map each node feature vector to pre-embedding vectors. Distinct fully connected layers were used for each node type. Two message passing layers were used each consisting of a dropout layer, a PReLU activation function, and a graph convolutional layer.

A custom heterogeneous graph attention layer was used using 1-head attention mostly following the structure of the original graph attention networks (Veličković et al., 2018), with the following modifications: 1) we applied fully connected layers with batch normalization to the node embeddings and the neighbor embeddings,and 2) we aggregated neighbor embeddings using the attention mechanism and concatenated the aggregated embedding to the current node’s embedding. This is then passed into a fully connected layer that reduces this down to a single output embedding followed by a batch normalization operation. Equation (1) shows our message passing function.

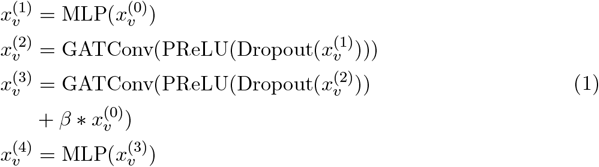

and Equation (2) shows the GATConv update function

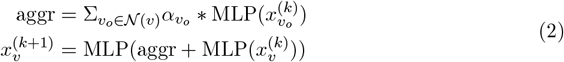

Where 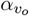 is the 1 head GAT attention score calculated for *v*_*o*_, 𝒩(*v*) is neighbors of *v*, and 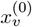 represents the node features of node *v*. The final predictions on existence of an ‘ordered-with’ edge *e*_*ij*_ between nodes 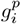 and 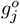 is inferred by concatenating their node embeddings and passing that through a fully connected two-layer perceptron, a batch normalization, a ReLU activation, and a final fully connected layer that outputs 2-dimensional logit vectors that are converted to final binary predictions using a softmax function. The formula for the link prediction head is as follows:

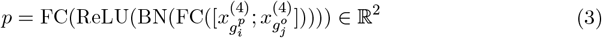

where BN refers to Batch Normalization and the first value corresponds to the probability that the edge exists and the second that it doesn’t.

## 3. Experimental Results

### 3.1. Endocrinology medical procedure recommendation

We used transductive disjoint training with a 1:4 positive:negative sampling implemented using PyG (Fey and Lenssen, 2019; pyg). Randomized cross validation with a set of 30 randomly selected hyper-parameter sets were used to tune the model. The final model uses Adam optimizer, dropout of 0.2, GAT convolutional layer, skip connection, and learning rate, hidden size, embedding size, and pre-embedding size are 1e-3, 64, 128 and 64, respectively. The model was tested on predictions made on all ‘ordered-with’ edges between a patient and an order placed during specialty visit and on an unseen test set consisting of 1, 321 patients.

Table 4 compares prediction results of our proposed GNN model with the baselines presented by Noshad et al. (2021) including fully connected multi-layer neural network (Diagnostic Model), a collaborative filtering auto-encoder (AE), singular value decomposition (SVD), probabilistic matrix factorization (PMF), an aggregate neural networks (Aggregated ANN), and an ensemble model (Ensemble Model) that uses a multi-layer neural network to combine the outputs of the diagnostic model, the collaborative filtering auto-encoder and the specialists’ identifiers as a separate input signal.

**Table 4:** Performance of endocrinologist procedure order prediction models.

| Model | AUC<br>ROC | P@R<br>0.50 | P@R<br>0.40 | P@R<br>0.30 |
| --- | --- | --- | --- | --- |
| Diagnostic Model | 0.65 | 0.33 | 0.42 | 0.46 |
| AE | 0.73 | 0.23 | 0.33 | 0.49 |
| PMF | 0.62 | 0.22 | 0.31 | 0.43 |
| SVD | 0.74 | 0.23 | 0.33 | 0.50 |
| Aggregated NN | 0.73 | 0.31 | 0.41 | 0.53 |
| Ensemble Model | 0.80 | 0.37 | 0.47 | 0.57 |
| GNN | 0.88 | 0.42 | 0.49 | 0.57 |

Our proposed model can predict endocrinology specialty procedure orders for the new patient referrals more effectively (ROC-AUC=0.88) compared to all models evaluated by Noshad et al. (2021) (best ROC-AUC of the baselines = 0.80). Further, our model showed significantly higher precision at recalls 0.5, 0.4 and 0.3 compared to all baseline models. Note, we used the same data as the data that were used in Noshad et al. (2021) except we removed features related to the specialists that patients were referred to, because although incorporating specialists’ information in the model can lead to even higher accuracy, the information on specific specialists in the clinic can add bias to the model.

Further, we compared our proposed GNN model with clinical checklist for endocrinology procedure recommendation. The proposed recommender algorithm approach provides medical procedure recommendations for endocrinology referrals more effectively than manual endocrinology checklists (recommender: precision=0.60, recall=0.27, F1-score=0.37) vs. (checklist: precision=0.16, recall=0.28, F1-score=0.20).

### 3.2. Hematology medical procedure recommendation

We compared our proposed GNN model’s performance in hematology procedure order recommendation with classical neural network and collaborative filtering based methods described in the previous section including a fully connected multi-layer neural network (Diagnostic Model), collaborative filtering auto-encoder (AE), singular value decomposition (SVD), probabilistic matrix factorization (PMF), aggregated neural networks (ANN), and an Ensemble Model combining the diagnostic model and the collaborative filtering autoencoder. Model tuning follows a randomized cross validation method similar to the model trained for the endocrinology cohort. Our optimized GNN model has GAT convolutions and the hidden size, embedding size, pre-embedding size and learning rate are 32, 128, 32, 1e-3, respectively. All models are tested using an unseen test set including 603 patients. The comparison results are presented in Table 5. Our proposed GNN model predicts procedures ordered during patients first visit with hematology clinic at least 5% more effectively in terms of ROC-AUC (ROC-AUC=0.84) compared to all baseline models. Further, the proposed model has higher precision at recalls 0.50 (precision = 0.41), 0.40 (precision = 0.44), and 0.30 (precision = 0.45) compared to the baselines.

**Table 5:** Performance of the models on medical procedure order recommendation for new referrals to hematology department.

| **Model** | **AUC**<br>**ROC** | **P@R**<br>**0.50** | **P@R**<br>**0.40** | **P@R**<br>**0.30** |
| --- | --- | --- | --- | --- |
| Diagnostic Model | 0.60 | 0.18 | 0.19 | 0.21 |
| AE | 0.49 | 0.17 | 0.22 | 0.33 |
| PMF | 0.38 | 0.10 | 0.16 | 0.22 |
| SVD | 0.74 | 0.38 | 0.43 | 0.46 |
| Aggregated NN | 0.64 | 0.24 | 0.27 | 0.32 |
| Ensemble Model | 0.79 | 0.40 | 0.41 | 0.41 |
| CR-GNN | 0.84 | 0.41 | 0.44 | 0.45 |

Further, we compared our proposed GNN model with the clinical checklist in active use by the healthcare system’s electronic consultation program. This checklist was produced by clinical committees in the health system to guide primary care providers when creating virtual consultations for XXXX hematology department. The checklist is offered for referral diagnoses including anemia, isolated erythrocytosis, elevated ferritin, isolated leukocytosis, isolated leukopenia, mgus, thrombocytopenia, thrombocytosis, and VTE/thrombophilia.

We compared the prediction performance of the proposed model with XXXX hematology electronic consultation checklist. The GNN recommender algorithm approaches provide medical procedure recommendations for hematology referrals more effectively than manual clinical checklists in terms of precision and F1-score (GNN recommender: precision=0.44, recall=0.38, F1-score=0.41) vs. (checklist: precision=0.27, recall=0.71, F1-score=0.39). We extracted referral diagnoses for the patients in our testing set and used the clinical checklist to predict the procedures they will need and compared these guideline suggestions against the procedures actually ordered during patients’ first visits at the hematology clinic as the ground truth. Utilizing the clinical checklist is not possible for many patients because the guideline doesn’t cover all referral diagnoses in the data. As a result, the test set for guideline is a subset of our original test set including 315 patients, and these results thus overestimate the performance of the guideline checklists given that they would not perform at all in the cases without a clear matching referral diagnosis that our recommender algorithm models are always able to adapt to.

We further explored the GNN model and hematologists behaviours for an example subset of the patients in our testing set who were referred to the hematology clinic with an anemia diagnosis (124 patients) as the most common referral diagnosis in our data. Figure 3 shows the top six procedures that were most frequently ordered by hematologists which naturally overlaps with the six procedures provided in the hematology checklist. A list of six procedures that were commonly recommended by our GNN model is also shown in Figure 3. Model’s predictions overlap with both the hematology checklist suggestions (vitamin B12, ferritin, transferrin saturation, CMP, CBC with differential, and reticulocyte count) as well as the commonly ordered procedures by hematologists including ferritin, transferrin saturation, metabolic panel, comprehensive, vitamin B12, reticulocyte count automated, and CBC with diff and slide review. Although, given the high accuracy of the model this was expected, this provides some explanations on the proposed model’s performance and shows that our GNN model’s recommendations are consistent with the guideline as well as the ground truth (the procedures ordered by the specialists). We further explored the target variables (procedures) where the model had highest true positives. The top 5 procedures (targets) which had the highest true positives (model recommended them correctly) include comprehensive metabolic panel, reticulocyte count, ferritin, transferrin saturation, and vitamin B12. Excluding the most commonly predicted targets by the model, procedures such as CBC with differential, transferrin saturation, and haptoglobin had the highest false positive rates (model recommended but not actually ordered in subsequent specialist visits).

**Figure 3:**
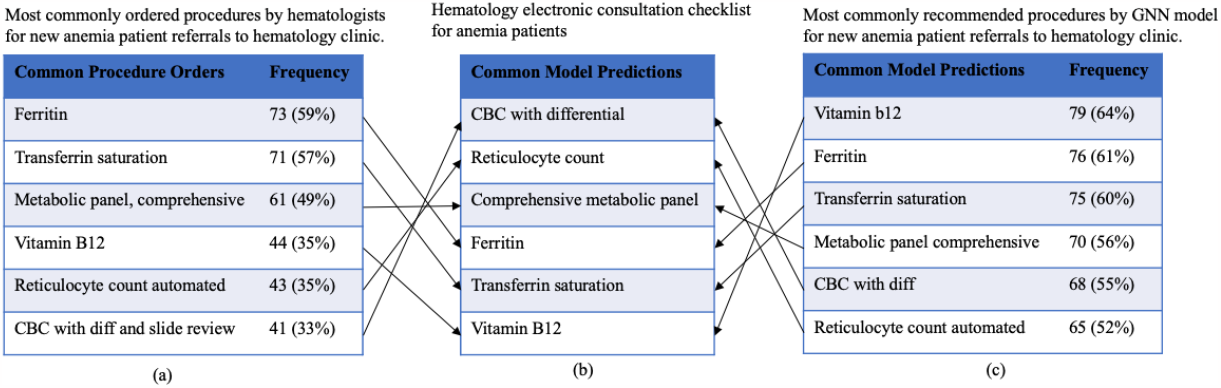
(a) Procedures commonly ordered by hematologists for new anemia patient referrals to the hematology clinic. (b) Procedures recommended by the hematology checklist for anemia referrals (c) GNN most common recommendations for new anemia patient referrals to the hematology clinic. Arrows connect similar procedures. Model predictions, hematology checklist, and procedures ordered by hematologists overlap indicates the consistency of the proposed GNN model decision making with the existing clinical checklist and the ground truth.

## 4. Discussion

In this study, we proposed a novel graph neural network based framework for medical procedure recommendation for specialty referral and virtual consultations. Models were trained and tested using new patients’ referrals to endocrinology and hematology clinics as two of the most common specialties with frequent consultation requests. Patients’ historical electronic health records were used to extract the predictors and the problem was models as a link prediction task in a heterogeneous graph structure. Proposed graph neural network based framework outperforms similar endocrinology and hematology medical procedure recommender algorithms in the literature in terms of ROC-AUC, precision, recall and f1-score. The GNN model outperforms endocrinology clinical checklists in terms of precision, recall and f1-score, and outperforms hematology clinical checklist in terms of precision and f1-score.

Clinical checklists are often limited to pre-defined set of diagnoses which precludes them from being used for a large group of patients with referral diagnoses outside of the scope of the guidelines. Our proposed AI models are end to end models that can analyze entire patient histories of EHR data and provide personalized recommendations. Using automated medical recommender tools could improve access to medical consultation guidance to patients by reducing the labor for clinicians and provide a consistent decision making support system for endocrinologists and hematologists and help them manage the ever-escalating complexity of electronic health records and medical guidelines. Primary care providers could also benefit from an order recommender system that suggests orders that sub-specialists might place, anticipating patient needs without time delays and space separation that both reduce access for vulnerable patient population. This study showed the opportunities and a pathway toward such an automated medical procedure recommender system.

Limitations in the study include that the models were built as an outpatient recommender system, but many of the features (notably hematology related chemotherapy support orders) were based in the inpatient setting. This may have implications for generalizability in settings where in-patient records are not as easily accessible. Therefore, available clinical checklists such as order set templates remain valuable for specialty care settings. It is worth noting that the current models were trained to recommend procedures ordered by specialists. However, these targets may include noise and human errors and may not necessarily be correct and accurate. More follow-up studies needed for outpatient based outcomes and reinforcement learning towards crowdsourced recommender items and literature evidence-based clinical practice checklist as well as association with patients outcomes

AI performance in personalized recommendation for medical procedures for endocrinology and hematology patients shows the potential of combining both AI and manual approaches to help primary care providers when referring patients for specialty care or requesting virtual consultation.

## Data Availability

The data include Stanford's healthcare patient and can not be publicly available.

### Appendix A. Variable Names

**Table A1:** List of diagnoses used as features in endocrinology cohort.

| Diagnosis name |
| --- |
| Diabetes mellitus Type I or II |
| Hypercalcemia |
| Hyperlipidemia |
| Hypothyroidism |
| Hyperthyroidism |
| Osteopenia |
| Osteoporosis |
| Thyroid cancer |
| Thyroid nodule |
| Obesity |

**Table A2:**
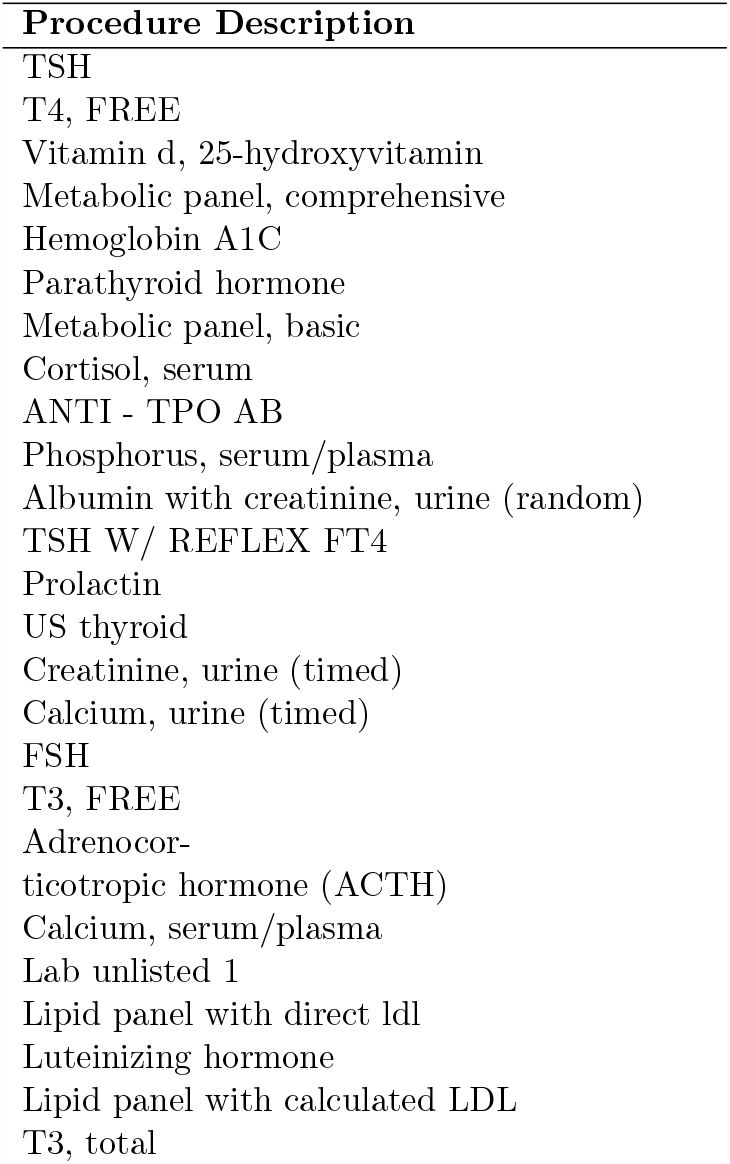
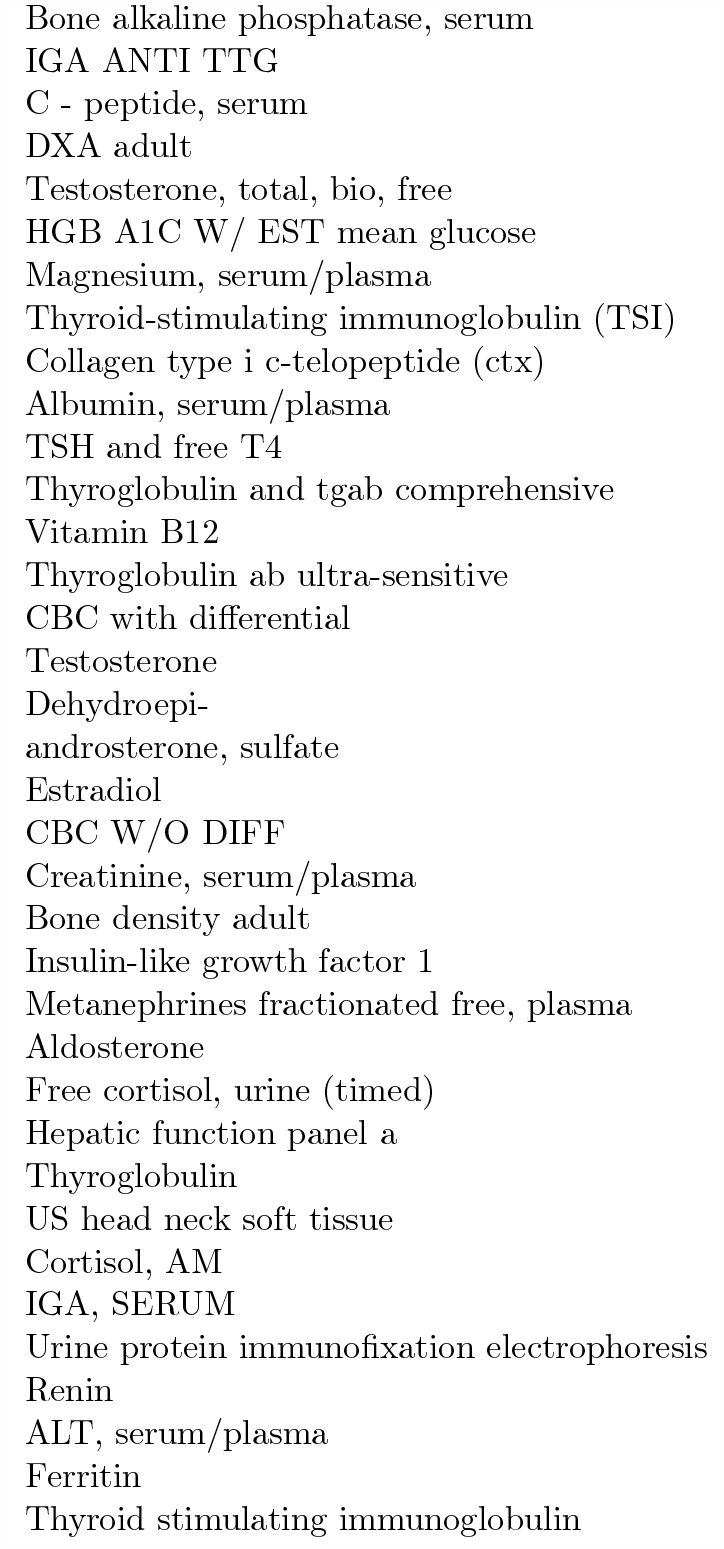
List of medical procedure features in endocrinology cohort.

**Table A3:**
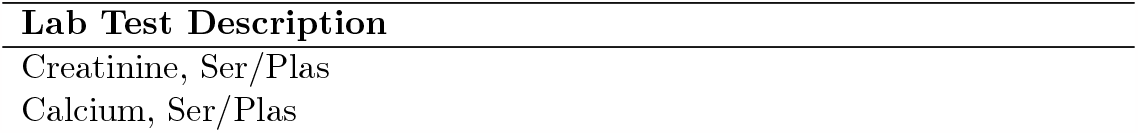
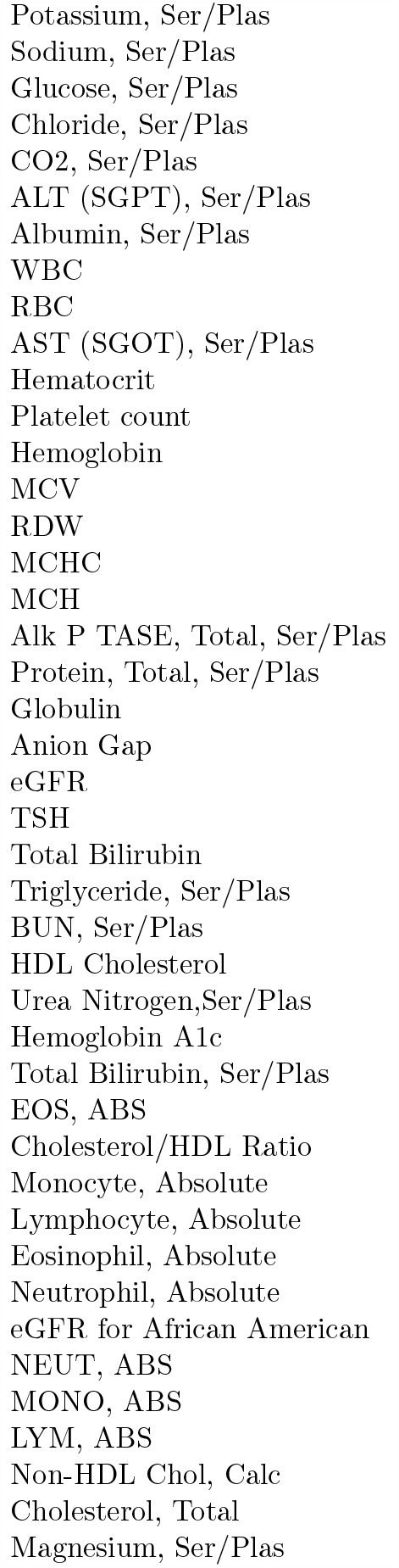
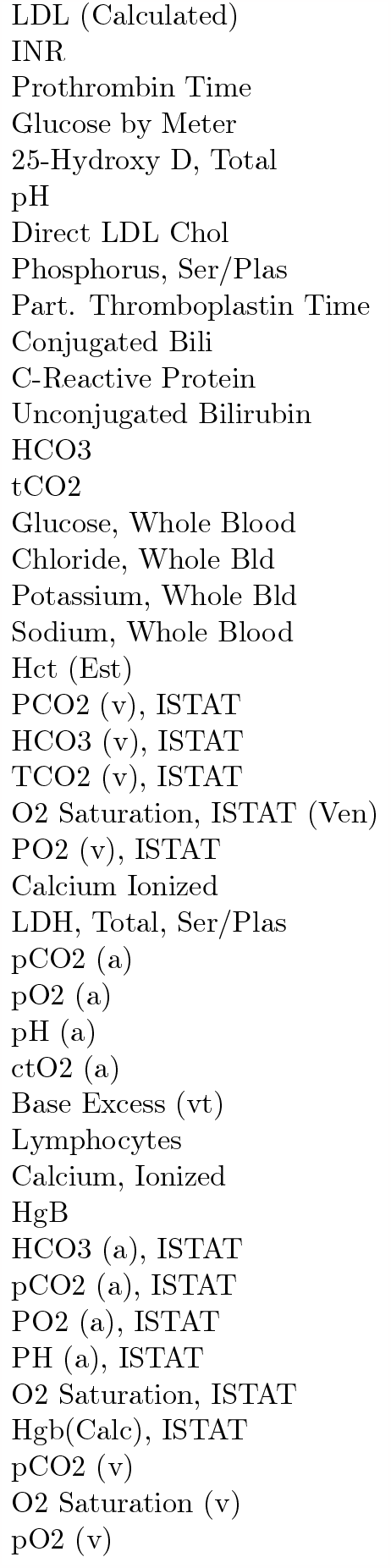
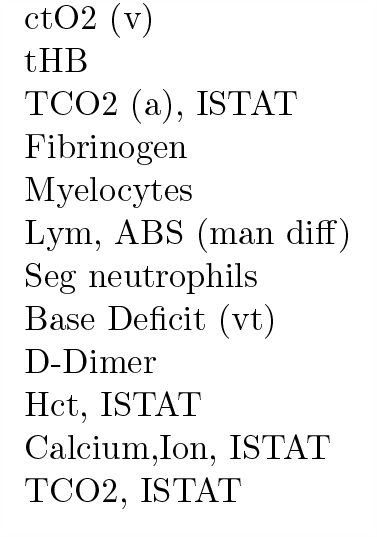
List of lab features in endocrinology cohort.

**Table A4:**
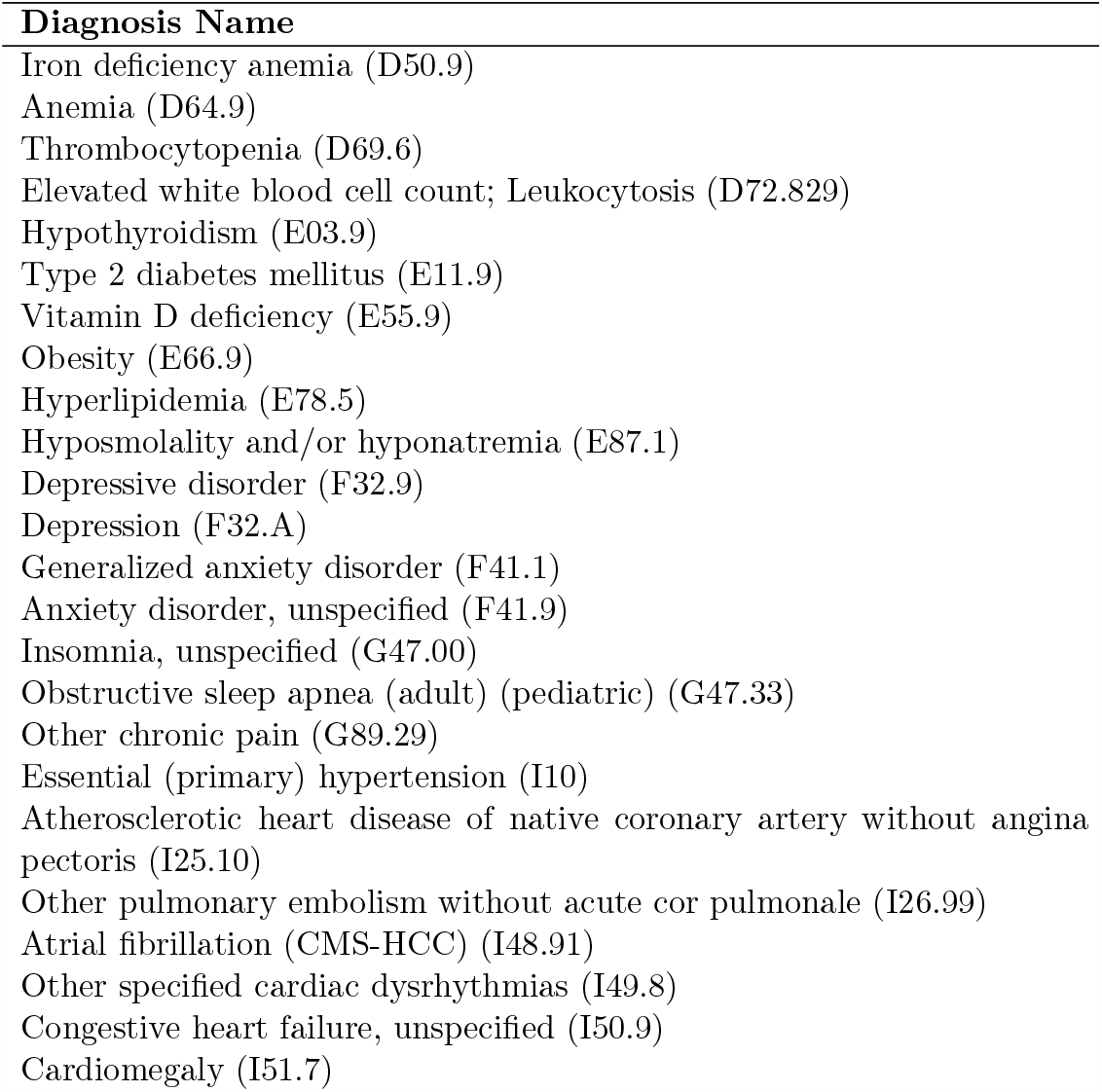
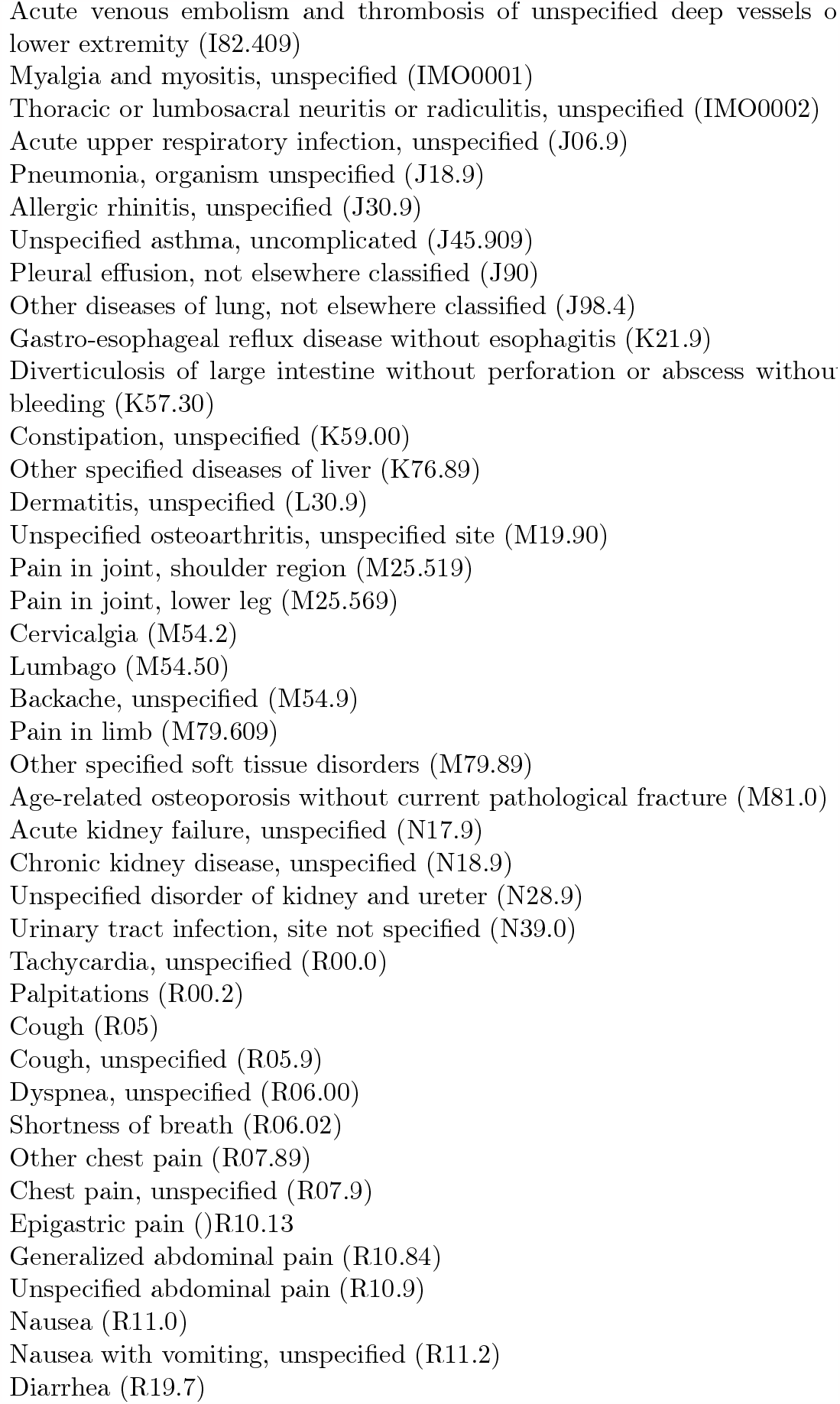
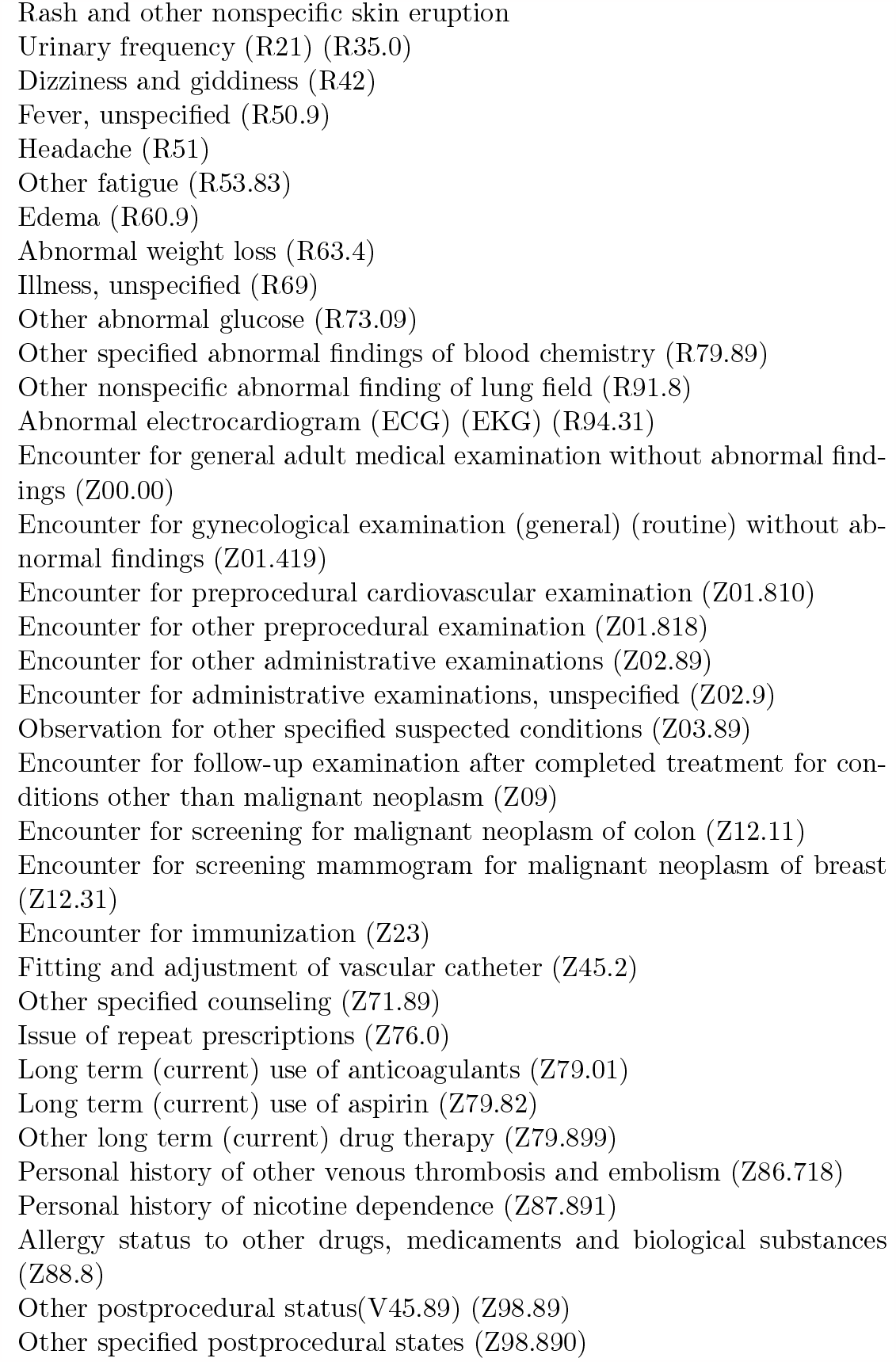
List of diagnoses features in hematology cohort.

| Diagnosis Name |
| --- |
| Iron deficiency anemia (D50.9) |
| Anemia (D64.9) |
| Thrombocytopenia (D69.6) |
| Elevated white blood cell count; Leukocytosis (D72.829) |
| Hypothyroidism (E03.9) |
| Type 2 diabetes mellitus (E11.9) |
| Vitamin D deficiency (E55.9) |
| Obesity (E66.9) |
| Hyperlipidemia (E78.5) |
| Hyposmolality and/or hyponatremia (E87.1) |
| Depressive disorder (F32.9) |
| Depression (F32.A) |
| Generalized anxiety disorder (F41.1) |
| Anxiety disorder, unspecified (F41.9) |
| Insomnia, unspecified (G47.00) |
| Obstructive sleep apnea (adult) (pediatric) (G47.33) |
| Other chronic pain (G89.29) |
| Essential (primary) hypertension (I10) |
| Atherosclerotic heart disease of native coronary artery without angina pectoris (I25.10) |
| Other pulmonary embolism without acute cor pulmonale (I26.99) |
| Atrial fibrillation (CMS-HCC) (I48.91) |
| Other specified cardiac dysrhythmias (I49.8) |
| Congestive heart failure, unspecified (I50.9) |
| Cardiomegaly (I51.7) |

Table A4: List of diagnoses features in hematology cohort.
| Diagnosis Name |
| --- |
| Acute venous embolism and thrombosis of unspecified deep vessels of lower extremity (I82.409) |
| Myalgia and myositis, unspecified (IMO0001) |
| Thoracic or lumbosacral neuritis or radiculitis, unspecified (IMO0002) |
| Acute upper respiratory infection, unspecified (J06.9) |
| Pneumonia, organism unspecified (J18.9) |
| Allergic rhinitis, unspecified (J30.9) |
| Unspecified asthma, uncomplicated (J45.909) |
| Pleural effusion, not elsewhere classified (J90) |
| Other diseases of lung, not elsewhere classified (J98.4) |
| Gastro-esophageal reflux disease without esophagitis (K21.9) |
| Diverticulosis of large intestine without perforation or abscess without bleeding (K57.30) |
| Constipation, unspecified (K59.00) |
| Other specified diseases of liver (K76.89) |
| Dermatitis, unspecified (L30.9) |
| Unspecified osteoarthritis, unspecified site (M19.90) |
| Pain in joint, shoulder region (M25.519) |
| Pain in joint, lower leg (M25.569) |
| Cervicalgia (M54.2) |
| Lumbago (M54.50) |
| Backache, unspecified (M54.9) |
| Pain in limb (M79.609) |
| Other specified soft tissue disorders (M79.89) |
| Age-related osteoporosis without current pathological fracture (M81.0) |
| Acute kidney failure, unspecified (N17.9) |
| Chronic kidney disease, unspecified (N18.9) |
| Unspecified disorder of kidney and ureter (N28.9) |
| Urinary tract infection, site not specified (N39.0) |
| Tachycardia, unspecified (R00.0) |
| Palpitations (R00.2) |
| Cough (R05) |
| Cough, unspecified (R05.9) |
| Dyspnea, unspecified (R06.00) |
| Shortness of breath (R06.02) |
| Other chest pain (R07.89) |
| Chest pain, unspecified (R07.9) |
| Epigastric pain (R10.13) |
| Generalized abdominal pain (R10.84) |
| Unspecified abdominal pain (R10.9) |
| Nausea (R11.0) |
| Nausea with vomiting, unspecified (R11.2) |
| Diarrhea (R19.7) |

Table A4: List of diagnoses features in hematology cohort.
| Diagnosis Name |
| --- |
| Rash and other nonspecific skin eruption |
| Urinary frequency (R21) (R35.0) |
| Dizziness and giddiness (R42) |
| Fever, unspecified (R50.9) |
| Headache (R51) |
| Other fatigue (R53.83) |
| Edema (R60.9) |
| Abnormal weight loss (R63.4) |
| Illness, unspecified (R69) |
| Other abnormal glucose (R73.09) |
| Other specified abnormal findings of blood chemistry (R79.89) |
| Other nonspecific abnormal finding of lung field (R91.8) |
| Abnormal electrocardiogram (ECG) (EKG) (R94.31) |
| Encounter for general adult medical examination without abnormal findings (Z00.00) |
| Encounter for gynecological examination (general) (routine) without abnormal findings (Z01.419) |
| Encounter for preprocedural cardiovascular examination (Z01.810) |
| Encounter for other preprocedural examination (Z01.818) |
| Encounter for other administrative examinations (Z02.89) |
| Encounter for administrative examinations, unspecified (Z02.9) |
| Observation for other specified suspected conditions (Z03.89) |
| Encounter for follow-up examination after completed treatment for conditions other than malignant neoplasm (Z09) |
| Encounter for screening for malignant neoplasm of colon (Z12.11) |
| Encounter for screening mammogram for malignant neoplasm of breast (Z12.31) |
| Encounter for immunization (Z23) |
| Fitting and adjustment of vascular catheter (Z45.2) |
| Other specified counseling (Z71.89) |
| Issue of repeat prescriptions (Z76.0) |
| Long term (current) use of anticoagulants (Z79.01) |
| Long term (current) use of aspirin (Z79.82) |
| Other long term (current) drug therapy (Z79.899) |
| Personal history of other venous thrombosis and embolism (Z86.718) |
| Personal history of nicotine dependence (Z87.891) |
| Allergy status to other drugs, medicaments and biological substances (Z88.8) |
| Other postprocedural status (V45.89) (Z98.89) |
| Other specified postprocedural states (Z98.890) |

**Table A5:**
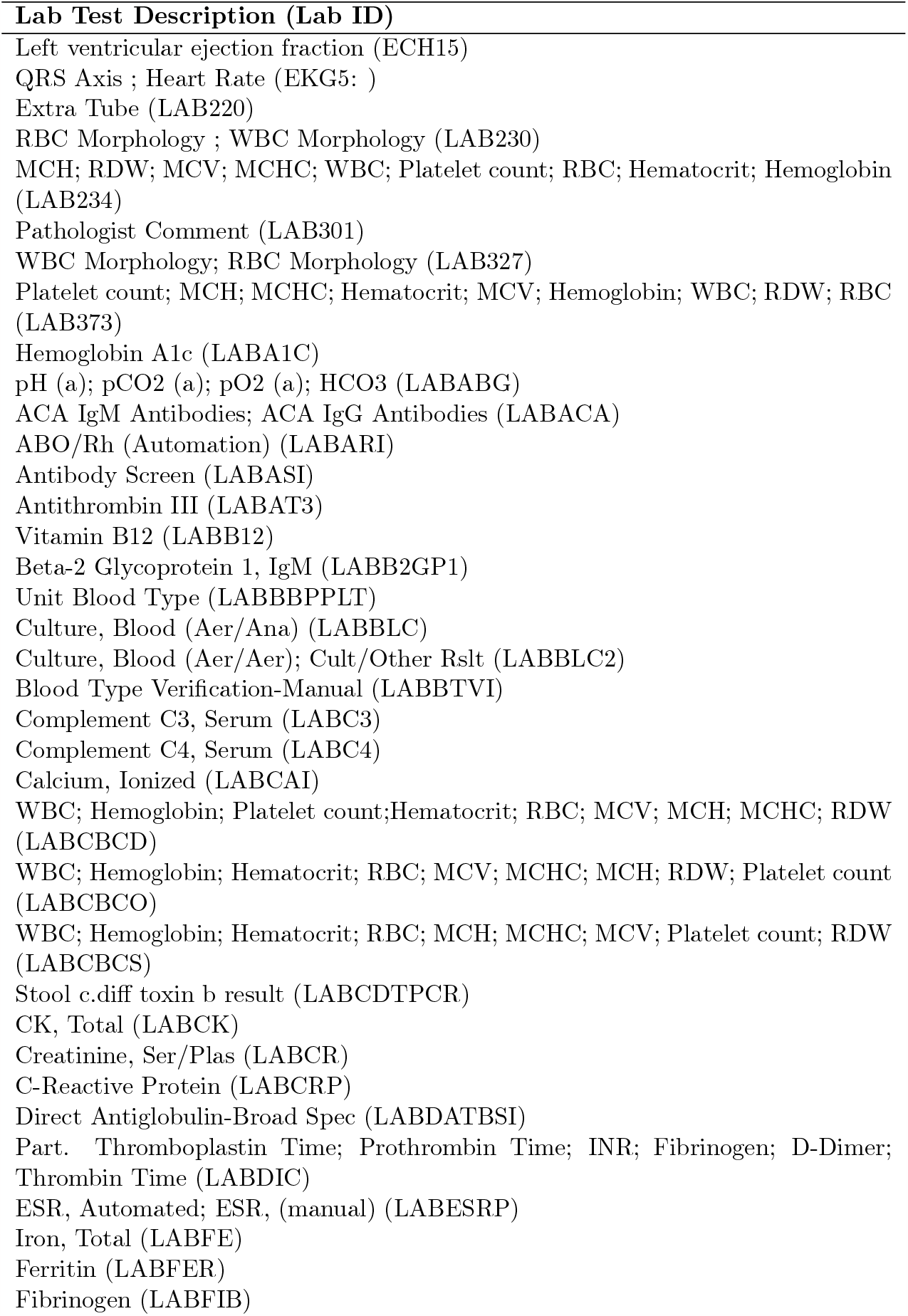
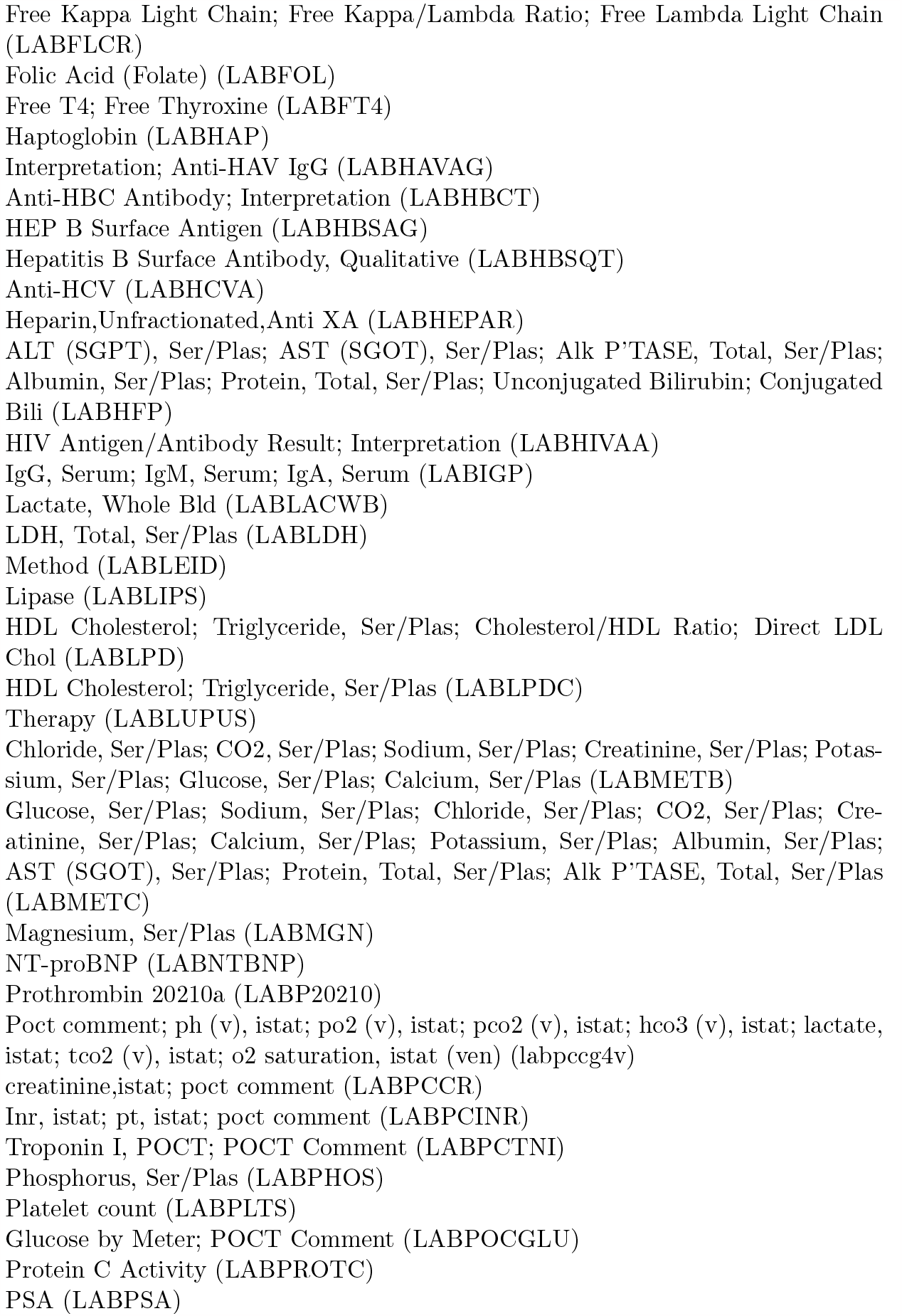
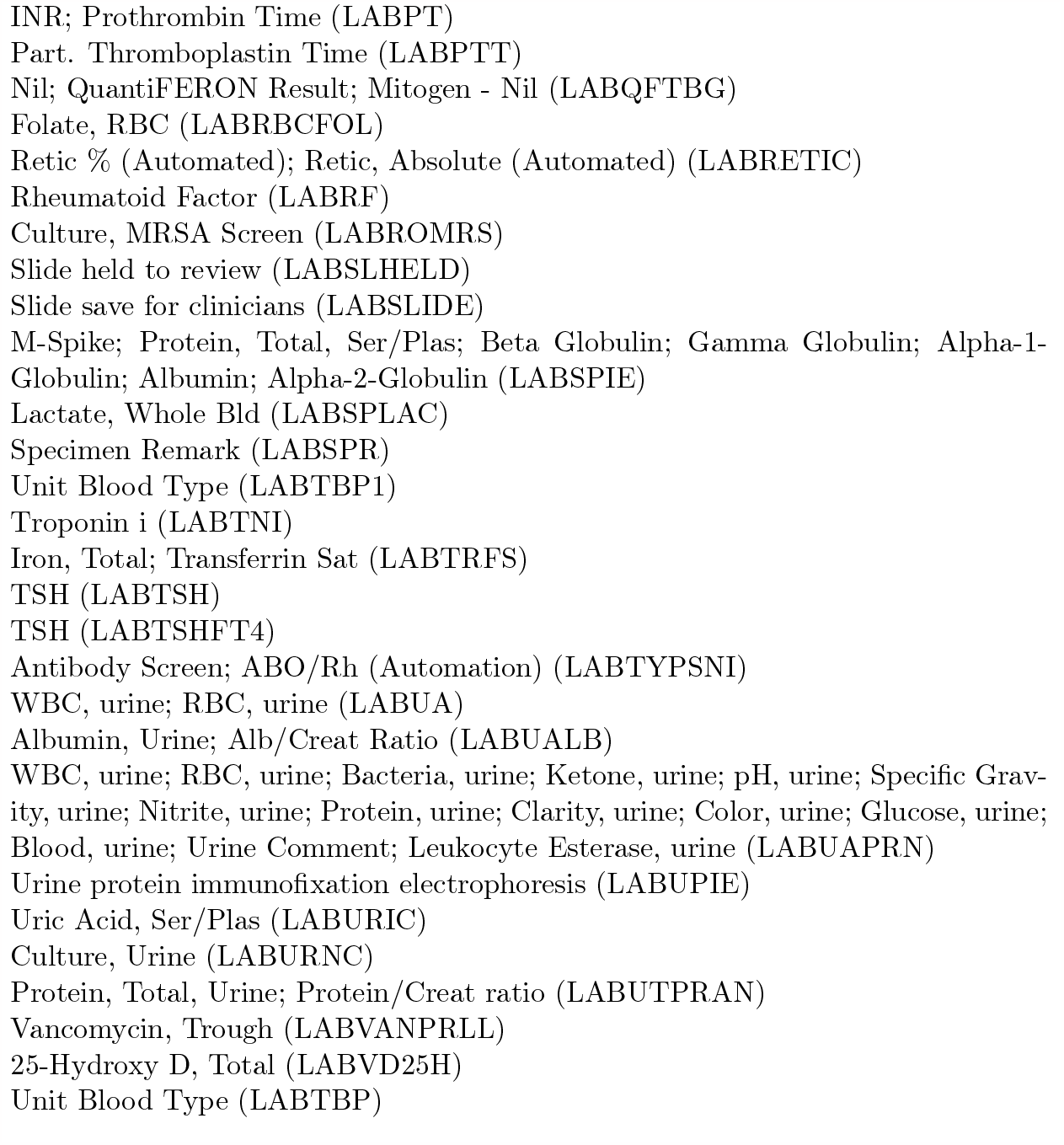
List of lab features for hematology cohort.

**Table A6:**
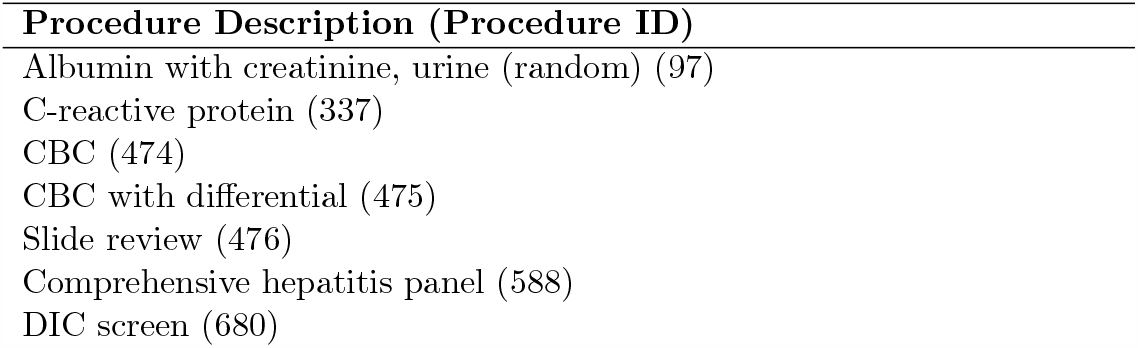
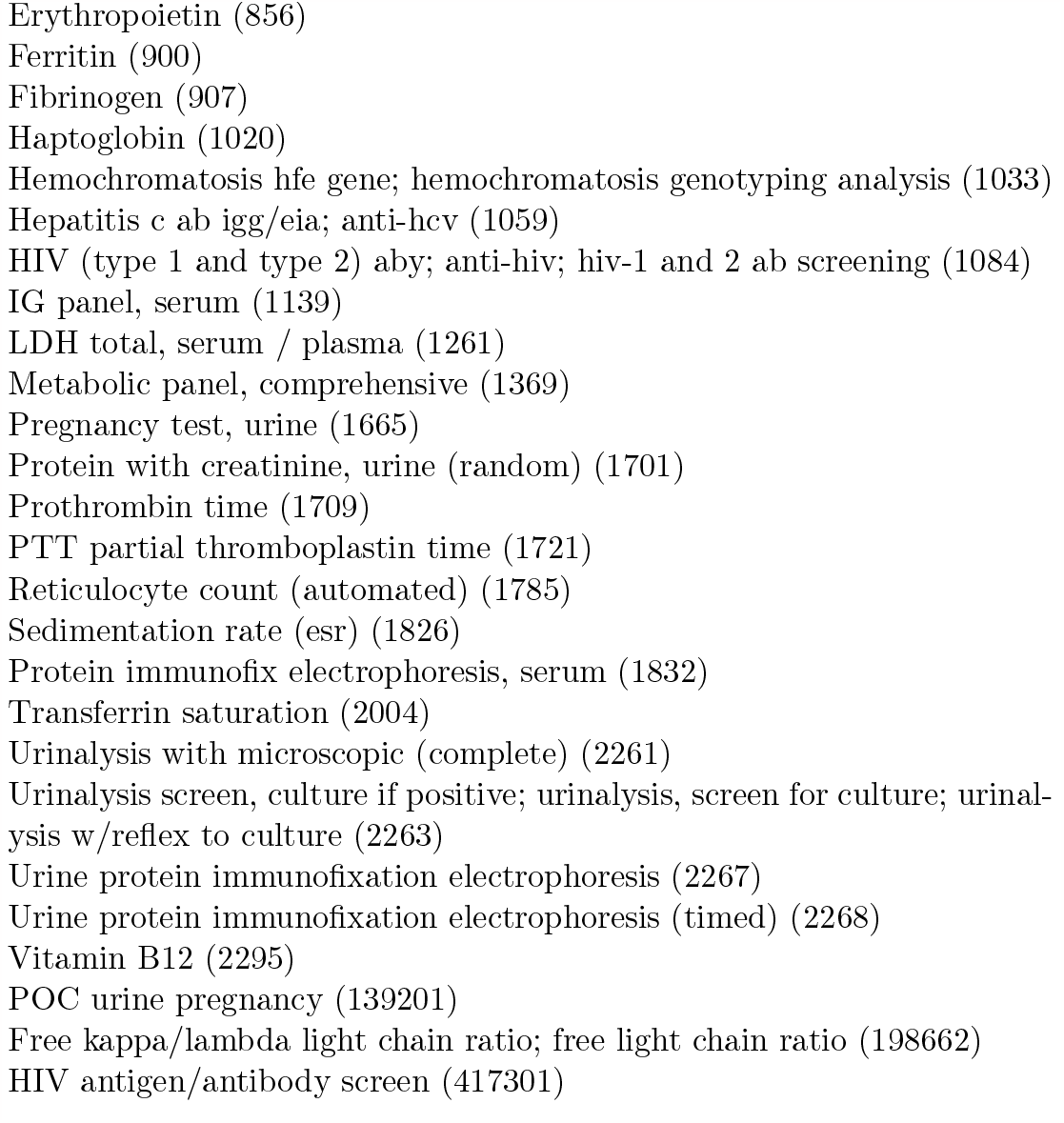
List of procedure features for hematology cohort.

| Procedure Description (Procedure ID) |
| --- |
| Albumin with creatinine, urine (random) (97) |
| C-reactive protein (337) |
| CBC (474) |
| CBC with differential (475) |
| Slide review (476) |
| Comprehensive hepatitis panel (588) |
| DIC screen (680) |

Table A6: List of procedure features for hematology cohort.
| **Procedure Description (Procedure ID)** |
| --- |
| Erythropoietin (856) |
| Ferritin (900) |
| Fibrinogen (907) |
| Haptoglobin (1020) |
| Hemochromatosis hfe gene; hemochromatosis genotyping analysis (1033) |
| Hepatitis c ab igg/eia; anti-hcv (1059) |
| HIV (type 1 and type 2) aby; anti-hiv; hiv-1 and 2 ab screening (1084) |
| IG panel, serum (1139) |
| LDH total, serum / plasma (1261) |
| Metabolic panel, comprehensive (1369) |
| Pregnancy test, urine (1665) |
| Protein with creatinine, urine (random) (1701) |
| Prothrombin time (1709) |
| PTT partial thromboplastin time (1721) |
| Reticulocyte count (automated) (1785) |
| Sedimentation rate (esr) (1826) |
| Protein immunofix electrophoresis, serum (1832) |
| Transferrin saturation (2004) |
| Urinalysis with microscopic (complete) (2261) |
| Urinalysis screen, culture if positive; urinalysis, screen for culture; urinalysis w/reflex to culture (2263) |
| Urine protein immunofixation electrophoresis (2267) |
| Urine protein immunofixation electrophoresis (timed) (2268) |
| Vitamin B12 (2295) |
| POC urine pregnancy (139201) |
| Free kappa/lambda light chain ratio; free light chain ratio (198662) |
| HIV antigen/antibody screen (417301) |

## Notes

### Competing Interest Statement

The authors have declared no competing interest.

### Funding Statement

This study did not receive any funding.

### Author Declarations

Stanford Institutional Review Board

### Summary of Updates

The manuscript has been extended to a full length paper. Models/results were updated. Please refer to/use/cite this revised version.

## References

PyG. https://www.pyg.org/. Accessed: 2022-09-31.

Eta S Berner and Tonya J La Lande. Overview of clinical decision support systems. Clinical decision support systems: Theory and practice, pages 1–17, 2016.

Tiffani J Bright, Anthony Wong, Ravi Dhurjati, Erin Bristow, Lori Bastian, Remy R Coeytaux, Gregory Samsa, Vic Hasselblad, John W Williams, Michael D Musty, et al. Effect of clinical decision-support systems: a systematic review. Annals of internal medicine, 157(1):29–43, 2012.

Jonathan H Chen, Tanya Podchiyska, and Russ B Altman. Orderrex: clinical order decision support and outcome predictions by data-mining electronic medical records. Journal of the American Medical Informatics Association, 23(2):339–348, 2016.

Jonathan Chiang, Andre Kumar, David Morales, Divya Saini, Jason Hom, Lisa Shieh, Mark Musen, Mary K Goldstein, and Jonathan H Chen. Physician usage and acceptance of a machine learning recommender system for simulated clinical order entry. AMIA Summits on Translational Science Proceedings, 2020:89, 2020.

Edward Choi, Mohammad Taha Bahadori, Le Song, Walter F Stewart, and Jimeng Sun. Gram: graph-based attention model for healthcare representation learning. In Proceedings of the 23rd ACM SIGKDD international conference on knowledge discovery and data mining, pages 787–795, 2017.

Edward Choi, Cao Xiao, Walter Stewart, and Jimeng Sun. Mime: Multilevel medical embedding of electronic health records for predictive healthcare. Advances in neural information processing systems, 31, 2018.

Edward Choi, Zhen Xu, Yujia Li, Michael Dusenberry, Gerardo Flores, Emily Xue, and Andrew Dai. Learning the graphical structure of electronic health records with graph convolutional transformer. In Proceedings of the AAAI conference on artificial intelligence, volume 34, pages 606–613, 2020.

Kathryn L Fantasia, Mary-Catherine Stockman, Zhihui Ju, Paola Ortega, Erika L Crable, Mari-Lynn Drainoni, Allan J Walkey, Megan Bergstrom, Katelyn O’Brien, and Devin Steenkamp. Professional continuous glucose monitoring and endocrinology econsult for adults with type 2 diabetes in primary care: results of a clinical pilot program. Journal of clinical & translational endocrinology, 24:100254, 2021.

Matthias Fey and Jan Eric Lenssen. Fast graph representation learning with pytorch geometric. arXiv preprint arXiv:1903.02428, 2019.

Will Hamilton, Zhitao Ying, and Jure Leskovec. Deepsnap. Advances in neural information processing systems, 30, 2017.

Haley S Hunter-Zinck, Jordan S Peck, Tania D Strout, and Stephan A Gaehde. Predicting emergency department orders with multilabel machine learning techniques and simulating effects on length of stay. Journal of the American Medical Informatics Association, 26 (12):1427–1436, 2019.

Wui Ip, Priya Prahalad, Jonathan Palma, Jonathan H Chen, et al. A data-driven algorithm to recommend initial clinical workup for outpatient specialty referral: Algorithm development and validation using electronic health record data and expert surveys. JMIR Medical Informatics, 10(3):e30104, 2022.

Erin Keely, Clare Liddy, and Amir Afkham. Utilization, benefits, and impact of an econsultation service across diverse specialties and primary care providers. Telemedicine and e-Health, 19(10):733–738, 2013.

Judy E Kim-Hwang, Alice Hm Chen, Douglas S Bell, David Guzman, Hal F Yee, and Margot B Kushel. Evaluating electronic referrals for specialty care at a public hospital. Journal of general internal medicine, 25(10):1123–1128, 2010.

Jeffrey Klann, Gunther Schadow, and J Michael McCoy. A recommendation algorithm for automating corollary order generation. In AMIA Annual Symposium Proceedings, volume 2009, page 333. American Medical Informatics Association, 2009.

Andre Kumar, Rachael C Aikens, Jason Hom, Lisa Shieh, Jonathan Chiang, David Morales, Divya Saini, Mark Musen, Michael Baiocchi, Russ Altman, et al. Orderrex clinical user testing: a randomized trial of recommender system decision support on simulated cases. Journal of the American Medical Informatics Association, 27(12):1850–1859, 2020.

Qimai Li, Zhichao Han, and Xiao-Ming Wu. Deeper insights into graph convolutional networks for semi-supervised learning. In Thirty-Second AAAI conference on artificial intelligence, 2018.

Ron C Li, Jason K Wang, Christopher Sharp, and Jonathan H Chen. When order sets do not align with clinician workflow: assessing practice patterns in the electronic health record. BMJ quality & safety, 28(12):987–996, 2019.

Clare Liddy, Margo S Rowan, Amir Afkham, Julie Maranger, and Erin Keely. Building access to specialist care through e-consultation. Open Medicine, 7(1):e1, 2013.

Zheng Liu, Xiaohan Li, Hao Peng, Lifang He, and S Yu Philip. Heterogeneous similarity graph neural network on electronic health records. In 2020 IEEE International Conference on Big Data (Big Data), pages 1196–1205. IEEE, 2020.

Zheng Liu, Xiaohan Li, Zeyu You, Tao Yang, Wei Fan, and Philip Yu. Medical triage chatbot diagnosis improvement via multi-relational hyperbolic graph neural network. In Proceedings of the 44th International ACM SIGIR Conference on Research and Development in Information Retrieval, pages 1965–1969, 2021.

Elizabeth A McGlynn, Steven M Asch, John Adams, Joan Keesey, Jennifer Hicks, Alison DeCristofaro, and Eve A Kerr. The quality of health care delivered to adults in the united states. New England journal of medicine, 348(26):2635–2645, 2003.

B Middleton, DF Sittig, and A Wright. Clinical decision support: a 25 year retrospective and a 25 year vision. Yearbook of medical informatics, 25(S 01):S103–S116, 2016.

Morteza Noshad, Ivana Jankovic, and Jonathan H Chen. Clinical recommender algorithms to simulate digital specialty consultations. In PACIFIC SYMPOSIUM ON BIOCOMPUTING 2022, pages 290–300. World Scientific, 2021.

Ted E Palen, David Price, Susan Shetterly, and Kristin B Wallace. Comparing virtual consults to traditional consults using an electronic health record: an observational case– control study. BMC medical informatics and decision making, 12(1):1–10, 2012.

Sungjin Park, Seongsu Bae, Jiho Kim, Tackeun Kim, and Edward Choi. Graph-text multimodal pre-training for medical representation learning. In Conference on Health, Inference, and Learning, pages 261–281. PMLR, 2022.

Julia C Prentice and Steven D Pizer. Delayed access to health care and mortality. Health services research, 42(2):644–662, 2007.

Barbara J Siepierski. Electronic consult experience: making health care more accessible and convenient for veterans. Fed Pract, 30(12):38–40, 2013.

Pierluigi Tricoci, Joseph M Allen, Judith M Kramer, Robert M Califf, and Sidney C Smith. Scientific evidence underlying the acc/aha clinical practice guidelines. Jama, 301(8): 831–841, 2009.

Petar Veličković, Guillem Cucurull, Arantxa Casanova, Adriana Romero, Pietro Liò, and Yoshua Bengio. Graph Attention Networks. International Conference on Learning Representations, 2018. URL https://openreview.net/forum?id=rJXMpikCZ.

Varsha G Vimalananda, Gouri Gupte, Siamak M Seraj, Jay Orlander, Dan Berlowitz, Benjamin G Fincke, and Steven R Simon. Electronic consultations (e-consults) to improve access to specialty care: a systematic review and narrative synthesis. Journal of telemedicine and telecare, 21(6):323–330, 2015.

Alina Vretinaris, Chuan Lei, Vasilis Efthymiou, Xiao Qin, and Fatma Özcan. Medical entity disambiguation using graph neural networks. In Proceedings of the 2021 International Conference on Management of Data, pages 2310–2318, 2021.

Jonathan X Wang, Delaney K Sullivan, Alex C Wells, and Jonathan H Chen. Clinicnet: Machine learning for personalized clinical order set recommendations. JAMIA open, 3 (2):216–224, 2020.

Paul Webster. Virtual health care in the era of covid-19. The Lancet, 395(10231):1180–1181, 2020.

Steffie Woolhandler and David U Himmelstein. The relationship of health insurance and mortality: is lack of insurance deadly? Annals of Internal Medicine, 167(6):424–431, 2017.

Tong Wu, Yunlong Wang, Yue Wang, Emily Zhao, and Yilian Yuan. Leveraging graph-based hierarchical medical entity embedding for healthcare applications. Scientific reports, 11 (1):1–13, 2021.

Yiye Zhang, Rema Padman, and James E Levin. Paving the cowpath: data-driven design of pediatric order sets. Journal of the American Medical Informatics Association, 21(e2): e304–e311, 2014.

Marinka Zitnik, Monica Agrawal, and Jure Leskovec. Modeling polypharmacy side effects with graph convolutional networks. Bioinformatics, 34(13):i457–i466, 2018.

